# Superspreading as a Regular Factor of the COVID-19 Pandemic: II. Quarantine Measures and the Second Wave

**DOI:** 10.1101/2020.08.14.20174557

**Authors:** Juri Dimaschko

## Abstract

Within the framework of a two-component model of the COVID-19 epidemic, taking into account the special role of *superspreaders*, we consider the impact of the recovery factor and quarantine measures on the course of the epidemic, as well as the possibility of a second wave of morbidity. It is assumed that there is no long-term immunity in asymptomatic *superspreaders* who have undergone the infection, and the emergence of long-term immunity in those who have undergone severe illness. It is shown that, under these assumptions, the relaxation of quarantine measures leads to the resumption of virus circulation among asymptomatic *superspreaders*. Depending on the characteristics of the quarantine, its removal may or may not lead to a renewed wave of daily morbidity. A criterion for the occurrence of repeated wave of morbidity is proposed based on the analysis of the final phase of the first wave. Based on this criterion, the repeated wave of the epidemic is predicted in New Zealand. A natural explanation is given for the decrease in lethality among the infected against the background of an absolute increase in their number.

## I. INTRODUCTION

In previous work [1], a two-component model of the COVID-19 epidemic was proposed. The model is based on the selection of two immunologically different groups of the population - *superspreaders* and *sensitive. Superspreaders* carry the infection without visible symptoms, so they spread it. *Sensitive*, having received an infection, fall ill, are isolated and therefore cannot spread it further. A few relevant examples have shown that the model adequately describes the course of the COVID-19 epidemic.

At the same time, the two-component model describes only the spread of the virus and does not consider the recovery processes. Further, it does not consider quarantine measures during the epidemic and the impact of these measures on the course of the epidemic itself. In this paper, we will include these factors in the two-component model and examine their impact on the final phase of the epidemic. In particular, the possibility of a second wave of the epidemic will be considered, and the conditions for its appearance will be determined.

The article is structured as follows.

In its second part, we consider the impact of recovery processes on the dynamics of the epidemic in the framework of the two-component model, as well as the impact of the quarantine as a factor affecting the spread rate. In this part, we introduce both factors into the dynamic equations of the two-component model and find an analytical solution under conditions of permanent quarantine.

The third part examines the 4 phases of the epidemic. In particular, the effect of lifting the quarantine in the final phase is being investigated. The effect is to re-increase the endemic equilibrium number of asymptomatic *superspreaders*. This, however, does not mean an automatic re-increase in the incidence, if the proportion of those who have been ill and who have received immunity among the *sensitive* is already large enough.

In the fourth section, we formulate a criterion for predicting the presence of a re-wave based on the analysis of the final segment of the first phase of the epidemic. Next, we compare the results obtained with the current course of the epidemic in a few countries and territories.

The final section summarizes the application of the model to the course of the current COVID-19 pandemic and provides a natural explanation for the decrease in mortality resulting from the two-component model.

## II. RECOVERY RATE AND PERMANENT QUARANTINE FACTOR

The dynamic equations of the two-component model [1] describe the change in the number of infected *sensitive* (*n*_1_ out of their total number *N*_1_) and the number of infected *superspreaders* (*n*_2_ out of their total number *N*_2_) over time:

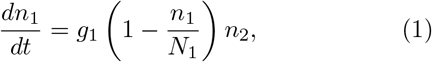

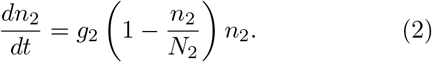

In this model, only *superspreaders* spread the infection, see Fig.1.

**Fig. 1:**
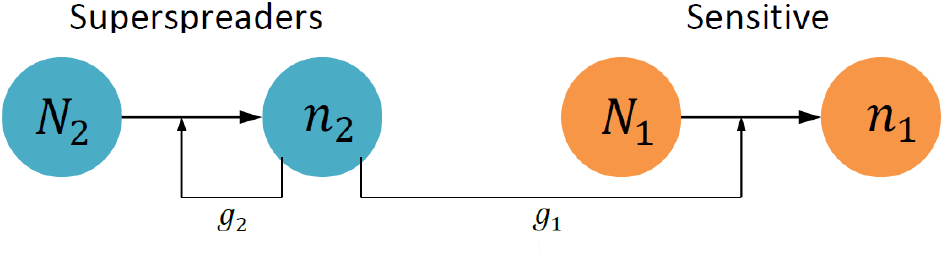
Scheme of the two-component epidemic model

Equations (1,2) take into account only the process of infection spread and do not take into account the recovery processes. In addition, they do not take into account the quarantine measures that have a direct impact on the spread rates of *g*_1_ and *g*_2_.

To consider this effect, we introduce the quarantine factor *Q*, which takes a value from 0 to 1. The effect of quarantine on the dynamics of the spread is reduced due to the same decrease in the spread rates *g*_1_ and *g*_2_:

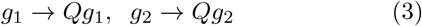

When considering the recovery processes, we will proceed from the assumption that after suffering a disease with severe symptoms, a person acquires absolute immunity. Thus, he is deprived of the opportunity to be re-infected and, from the point of view of the model, is no different from a simple patient - both during the illness and after it, a person from the *sensitive* group cannot spread the infection. Therefore, such a restoration does not affect the dynamics of the two-component model.

In contrast, recovery processes in asymptomatic infected individuals have a direct impact on the spread of the virus. We will assume that asymptomatic infected are deactivated with a recovery rate of γ. However, they do not acquire any lasting immunity and can be re-infected. This creates a circulation of infection among the *superspreaders*, leading to endemic equilibrium.

Considering the factors of quarantine *Q* and the recovery rate γ, the equations of the two-component model take the form

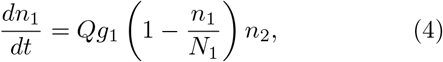

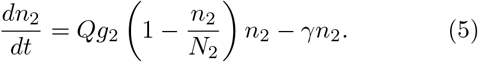

After switching to new variables

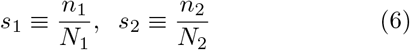

it is reduced to a simple form:

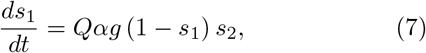

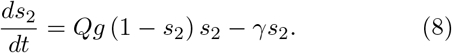

Here the value

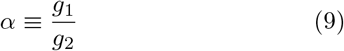

is the ratio of the spread rates *g*_1_ *≡ αg* and *g*_2_ *≡ g*. In the case of a long-time permanent quarantine, *Q* = const, this system of equations has an exact solution

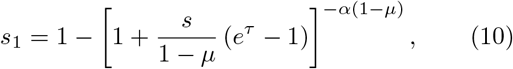

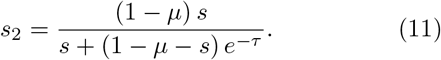

where *τ* = *Qgt* is dimensionless time and

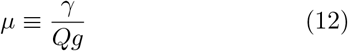

- dimensionless parameter of *superspreaders* deactivation rate. As in the absence of the quarantine (*Q* = 1) we have an epidemic occurs, the epidemic criterion is met (*μ* < 1). Consequently, the ratio of the recovery (*γ*) and spread (*g*) rates is also less than one:

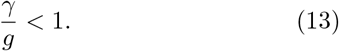

The found solution (10,11) meets the initial conditions

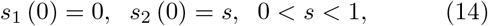

that is, at the beginning of the epidemic, there are already infected people, but no sick ones yet.

A stable endemic equilibrium among *superspreaders* is the state of

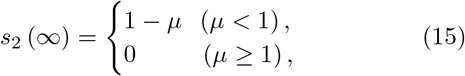

In accordance with (12,15), at a sufficiently small value of *Q*, i.e. with a sufficiently strict quarantine leading to the case of *μ* ≥ 1, *s*_2_ = 0 - all *superspreaders* are deactivated. In the opposite case of non-strict quarantine, when the value of the quarantine factor *Q* is not small enough and the parameter *μ* is less than one (*μ* < 1), an endemic equilibrium takes place with a certain proportion of active infected *superspreaders, s*_2_ > 0.

In such a solution the proportion of cases sensitive *s*_1_ continues to grow until it reaches the limit value *s*_1_ = 1. It is this condition that corresponds to the end of the epidemic and the vanishing of the daily incidench among *superspreaders* the infection continues, as at the beginning of the epidemic, to circulate at the stationary endemic level. Plots of the daily incidence *ds*_1_/*dt* according to the found solution (10, 11) for different values of the quarantine factor *Q* is shown in Fig. 2. As it should be, quarantine suppresses the amplitude of the disease wave. At the same time, it increases its duration.

**Fig. 2:**
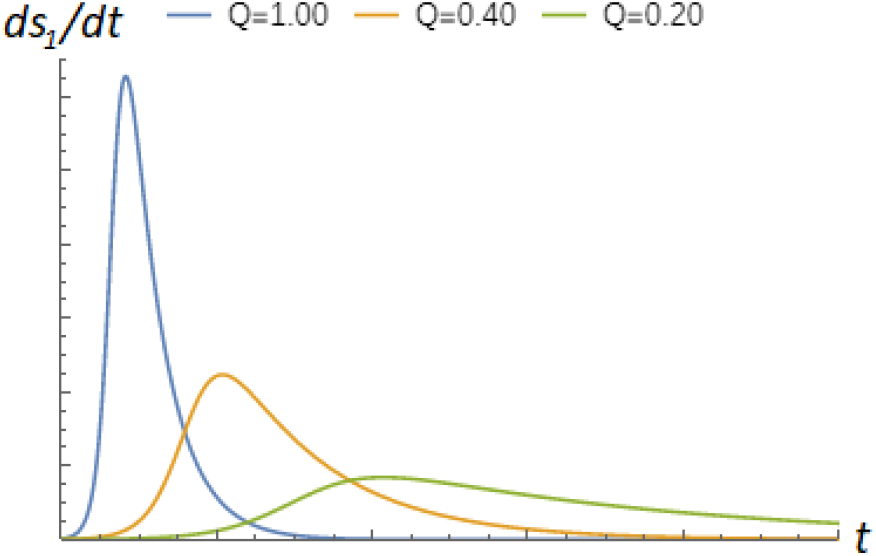
Plots of the daily incidence *ds*_1_*/dt* for different values of the quarantine factor *Q*. All plots correspond to *α* = 0.25. In the absence of the quarantine (*Q* = 1) the dimensionless recovery parameter *μ* = 0.1, in other two cases it is inverse proportional to the *Q*.

Thus, in the two-component model, the quarantine factor *Q* does not affect the reproductive number, which determines the exponential growth rate of the incidence, but rather the endemic equilibrium number of asymptomatic *superspreaders*. The daily incidence is directly proportional to this number. In addition, it is proportional to the proportion of those who do not recover among *sensitive*, which decreases during the epidemic. The end of the epidemic in this model does not correspond to the disappearance of infected *superspreaders*, but to the exhaustion of the number of still not infected *sensitive* individuals, which constitute only a relatively small part of the total population. The whole course of the epidemic in the two-component model is shown on a diagram in Fig.3.

**Fig. 3:**
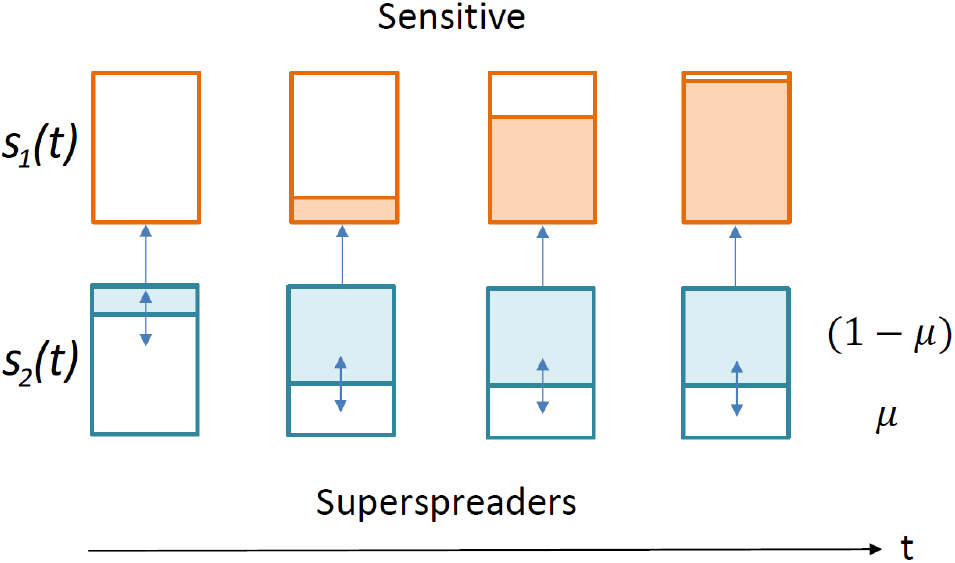
Course of an epidemic within the two-component model. Filled areas show share of infected *sensitive s*_1_(*t*) (orange) and *superspreaders s*_2_(*t*) (blue). When *t* → ∞, the share of infected *sensitive s*_1_(*t*) tends to 1, the share of infected *superspreaders s*_2_ (*t*) tends to the endemic equilibrium value of (1 − *μ*).

## III. VARIABLE QUARANTINE FACTOR AND THE SECOND WAVE

In the general case, the value of the quarantine factor *Q* is some function of time *Q*(*t*), determined by the sequence and severity of quarantine measures. A typical course of the function *Q*(*t*) is shown in Fig.4. Natural boundary conditions for it are

**Fig. 4:**
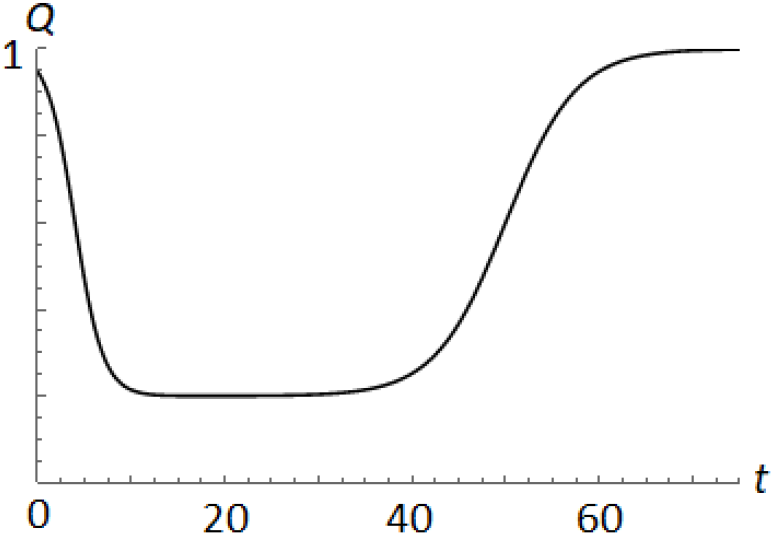
Quarantine factor *Q*(*t*), typical time dependence.

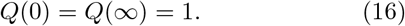

At the beginning of the epidemic, there is no quarantine and the quarantine factor is close to one, which corresponds to the boundary condition *Q*(0) = 1. As the epidemic develops, quarantine measures are taken, which are responsible for reducing the value of the *Q* factor. As the incidence decreases, the quarantine measures are removed, and the quarantine factor returns to the initial value. This corresponds to the second boundary condition *Q*(∞) = 1.

For an arbitrary dependence *Q*(*t*), the dynamic equations (7,8) have no exact solution. However, taking into account the boundary conditions (16), it turns out to be possible to carry out a qualitative analysis of the course of the epidemic corresponding to the real dependence *Q*(*t*). In this analysis, it is convenient to divide the entire course of the epidemic into 4 phases.

A) **The epidemic phase itself**. At the beginning of the epidemic, there is no quarantine,*Q* = 1. At the same time, the number of the *superspreaders* is growing exponentially, 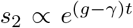. The daily incidence among *sensitives* is directly proportional to the number of *superspreaders ds*_1_*/dt* ∝*s*_2_ and therefore also grows exponentially.

B) **Quarantine start phase**. In this phase, the spread rates are suppressed by quarantine measures with a factor of *Q* < 1, the relative number of the *superspreaders* reaches the endemic equilibrium value *s*_2_ = 1 − *γ/*(*Qg*) and stops growing. Therefore, the daily incidence *ds*_1_ */dt*, which is directly proportional to the number of *superspreaders*, also stops growing. In this phase, the daily incidence reaches its maximum.

C) **Endemic phase**. In conditions of constant quarantine and, accordingly, an endemic constant number of the *superspreaders*, the daily incidence is gradually decreasing due to a decrease in the proportion of those who have not been cured among sensitive, (1 − *s*_1_). Ideally, by the end of this phase, this proportion is already small, 1 − *s*_1_ ≪ 1.

D) **Quarantine release phase**. After the quarantine is released, the equilibrium number of the *superspreaders* increases from the value *s*_2_ = 1 − *γ/*(*Qg*) established after the quarantine is turned on to the maximum possible value *s*_2_ = 1 − γ/g.

Thus, even if the endemic equilibrium number of the *superspreaders* was significantly reduced during quarantine, after the quarantine was lifted at the end of the epidemic, it again increases to its maximum value.

If the proportion of those who did not recover among the *sensitive* by the time the quarantine was lifted is already small, 1 − *s*_1_ ≪ 1, then this does not lead to a noticeable increase in the daily incidence at the end of the epidemic.

If this share has not yet managed to become sufficiently small, i.e. a significant part of the *sensitive*, then the release of quarantine leads to a noticeable increase in the daily incidence, i.e. to the second wave.

To illustrate the possibility of the emergence of the second wave, let us consider the course of the epidemic at different values of the quarantine factor *Q*, shown in Fig. 5. In the case of variable quarantine *Q*(*t*) we treat the quarantine factor *Q* as the minimal value of the function *Q*(*t*). This factor controls the endemic equilibrium number of the *superspreaders* during quarantine.

**Fig. 5.**
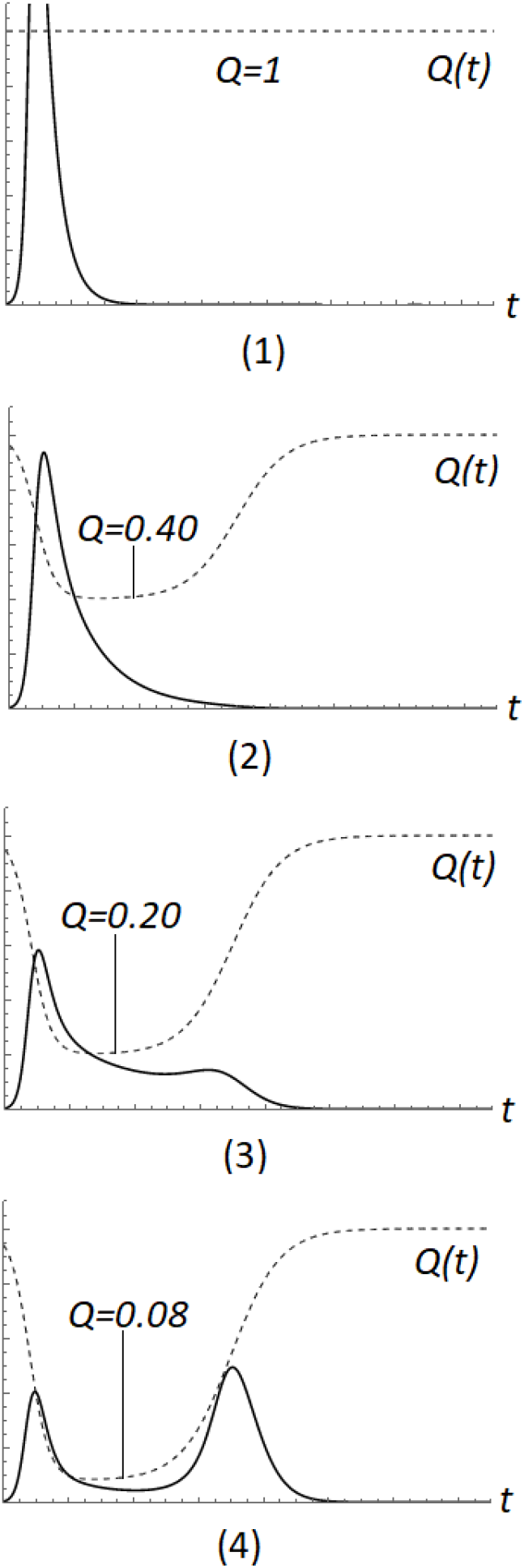
The appearance of the second wave with a decrease in the quarantine parameter *Q* (i.e., with an increase of the quarantine). The model parameters are taken as *α* = 0.25 and *μ* = 0.1. Sections A, B, C, D correspond to 4 different phases of the epidemic: A - the epidemic phase itself, B - the phase of the beginning of quarantine, C - the endemic phase, D - the phase of quarantine lifting. The dotted line shows the quarantine factor *Q*(*t*) as a function of time. solid - daily incidence. For comparison, the course of morbidity is shown in the complete absence of quarantine measures, *Q*=1. In this case, the maximum incidence would exceed the maximum scale of the graph by about 1.5 times. The minimum value of the quarantine factor *Q*=0.40 conditionally corresponds to moderate quarantine measures, *Q*=0.20 - strict, *Q*=0.08 - extremely strict.

At *Q* = 1 (absence of the quarantine), *Q* = 0.40 and *Q* = 0.20 (soft or moderately strict quarantine), the equilibrium endemic number of the *superspreaders* differs from zero. In these three cases, *superspreaders* continue to spread the infection during quarantine, and the incidence decreases relatively slowly due to a decrease in the number of *sensitive* who have not been ill. As can be seen from the first three graphs, here the release of quarantine after a significant decrease in the incidence does not lead to the appearance of a noticeable second wave, since by this moment the vast majority of the *sensitive* have already been ill.

If *Q* = 0.08 (very strict quarantine), as shown in the fourth graph, then the endemic equilibrium number of *superspreaders* during quarantine becomes zero. On the one hand, this leads to a rapid zero morbidity during quarantine. However, it is for this reason that most *sensitive* do not have time to get sick during quarantine. After the quarantine is released, the number of the *superspreaders* returns to the former endemic equilibrium non-zero value, and a new wave of infected people appears among the *sensitive*. It represents the second (residual) wave of morbidity

Thus excessively strict quarantine that deactivates all carriers of the virus is harmful. The reason for this is that in the absence of the virus, the *sensitive* are completely deprived of the opportunity to acquire immunity during the course of the disease. When active infected *superspreaders* reappear after the quarantine has been lifted, this still inevitably leads to the infection of the *sensitive*, deprived of immunity, to the appearance of a second wave of the epidemic and, thereby, to an increase in its duration.

## IV. SECOND WAVE CRITERION AND ANALYSIS OF THE REAL COURSES OF THE EPIDEMIC

Since the severity of quarantine can be assessed only by its consequences, it seems reasonable to find a criterion for the possibility of a second wave after quarantine is removed (corresponds to the phase D in Fig.5) based on the analysis of the current course of the epidemic, i.e. before the quarantine was lifted (corresponds to the start quarantine phase B and endemic phase C in Fig. 5).

The forecast of the second wave will be based on the course of the epidemic immediately before and after the first maximum. In accordance with the exact solution (10,11), before reaching the first maximum, the increase in the incidence rate under quarantine conditions with factor *Q* occurs with an exponential rate 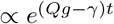. After reaching the first maximum, the incidence rate decreases exponentially 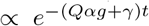. The ratio of the increment of increase and decrement of decrease is the value

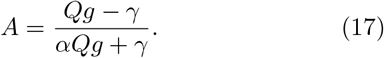

In the epidemic limit γ*/*(*Qg*) ≪ 1, when the recovery constant can be neglected, this ratio is equal to 1/*α*. In [1], we showed, using the examples of a number of countries, that this value has a numerical value of

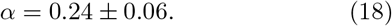

This means that for finite *γ*, the value of *A* can not exceed 1/*α* ≈ 4. The lower the quarantine factor *Q*, that is the stricter the quarantine, the smaller the value of *A*. In accordance with Fig.5, the lower *Q*, the more likely the second wave. Consequently, the small value of *A* ≪ 4 indicates appearance of the second wave.

Examples of this kind are Israel (*A*=1.1) and Serbia (*A*=0.9), where there is a significant second wave. The course of the epidemic in these countries is shown in Fig.6. The value of the critical parameter *A* is determined by the section of the graph at half the maximum height and the projection of the top of the graph onto this section. The value of parameter *A* in this and subsequent graphs is estimated as the ratio of the lengths of the right and left segments of this section.

**Fig. 6.**
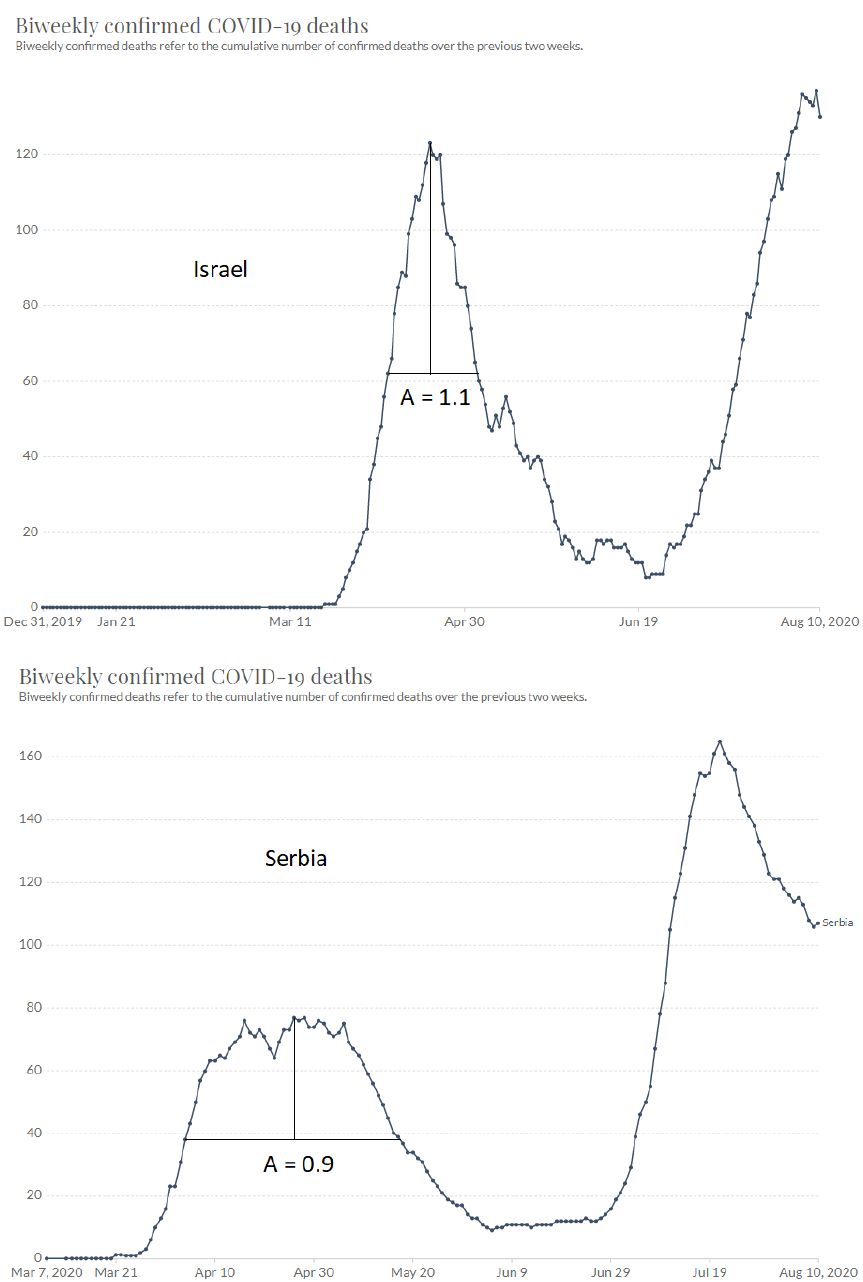
The course of the COVID-19 epidemic (deaths in the 14 days preceding the current one) in Israel and Serbia.

A very interesting situation shown in Fig.7 is observed in Australia and New Zealand. The first peak of the epidemics in both neighbouring countries is similar, observed in the same time and satisfies the second wave criterion: *A*=0.9 in Australia and *A*=1.0 in New Zealand. In Australia we already observe the second wave with maximum about 500 cases per day. The value of *A*=1.0 indicates the emergence of a similar second wave in New Zealand in the near future.

**Fig. 7.**
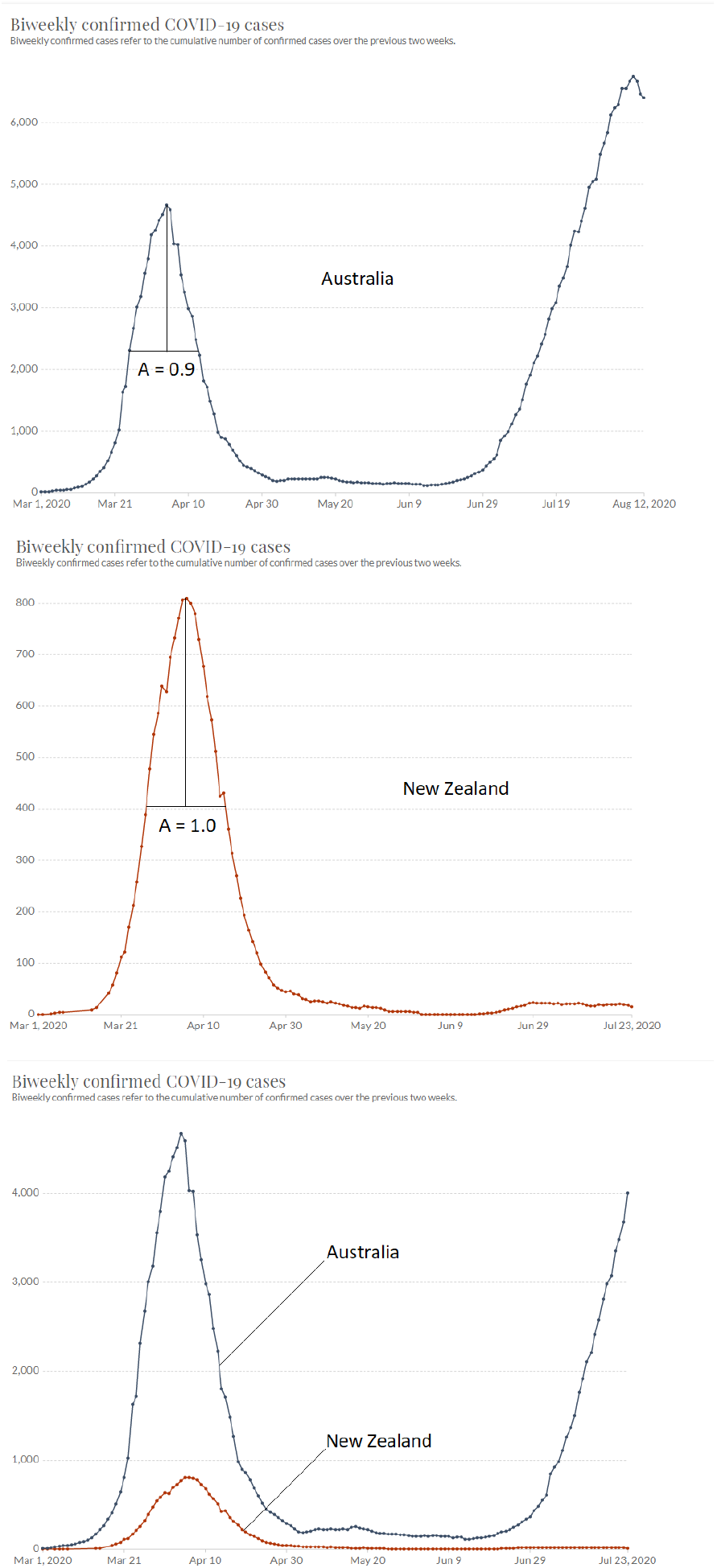
The course of the COVID-19 epidemic (cases in the 14 days preceding the current one) in Australia and New Zealand. The two-component model predicts the second wave in New Zealand.

Counterexamples of countries with moderate values of A are Germany (*A*=1.5) and Italy (*A*=2.5), where the signs of the second wave are very weak. The course of the epidemic in these countries is shown in Fig.8.

**Fig. 8.**
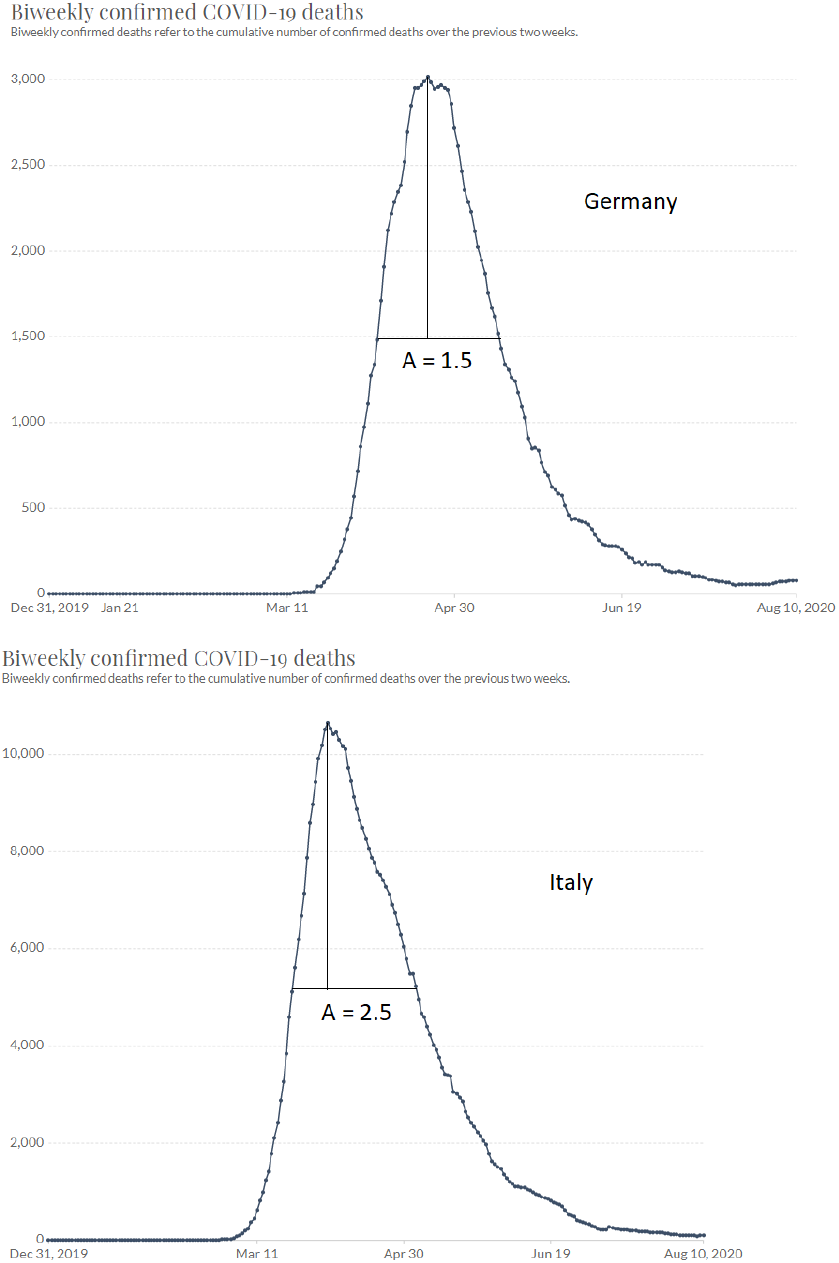
The course of the COVID-19 epidemic (deaths in the 14 days preceding the current one) in Germany and Italy.

In addition to the excessively strict quarantine, the reason for the appearance of the second wave can obviously be its premature weakening. In this case, the endemic equilibrium number of *superspreaders* increases against the background of a significant proportion of *sensitive* individuals who have not recovered and have not yet received immunity. This will lead to an immediate increase in the incidence, as appears to have happened in Iran and the United States. The course of the epidemic in these countries is shown in Fig.9.

**Fig. 9.**
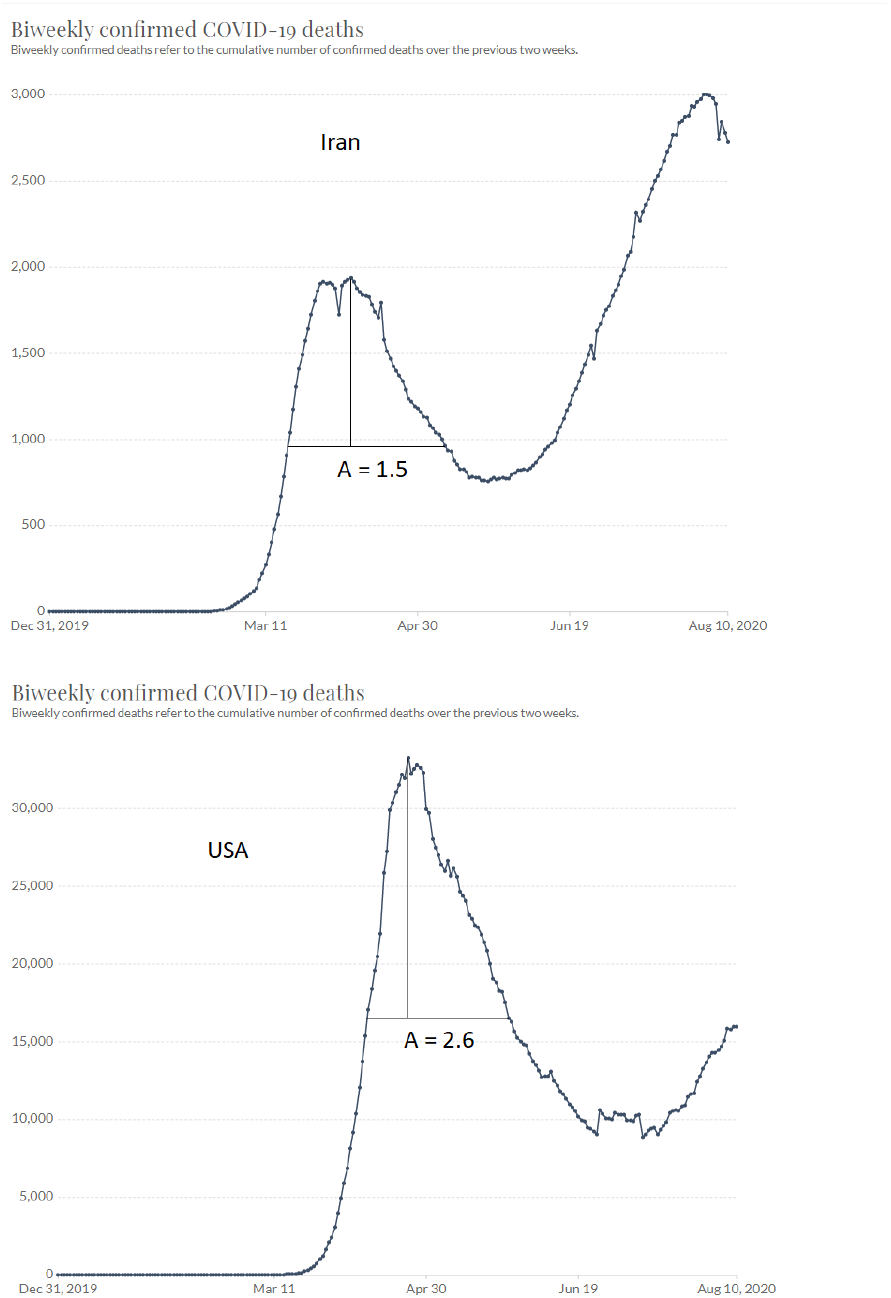
The course of the COVID-19 epidemic (deaths in the 14 days preceding the current one) in Iran and USA.

Note that in terms of the criterion of severity of quarantine, Iran is equivalent to Germany, and the United States is equivalent to Italy (Fig. 8). This indicates that in both cases, extending the quarantine until the end of the first wave would have avoided the appearance of a second wave.

## V. CONCLUSIONS AND DISCUSSION

Thus, the two-component model not only provides an adequate description of the course of the COVID-19 epidemic, but also allows one to assess the impact of the severity and duration of quarantine measures on the course of the epidemic. It results a simple criterion for predicting the possibility of a second wave of the epidemic after the quarantine is weakened. Based on this criterion, we can **predict** the repeated wave of the epidemic in New Zealand.

Further, the two-component model provides a natural explanation for the observed decrease in mortality among those infected. It is important to understand that, due to the individual differences in the immune response, the concepts of “infected” and “sick” are not equivalent: infection leads to disease only in *sensitive* and does not lead to disease in *superspreaders*. The increase in the number of infected in the final phase of the epidemic may largely occur not due to an increase in real morbidity, but due to an increase in the number of asymptomatically infected *superspreaders* found during testing. Since those who have recovered from the *sensitive* acquire immunity, an increasing proportion of those infected are *superspreaders*. That is why, according to the two-component model, there is a decrease in mortality among the full array of infected people, which consists of two parts: sick - infected sensitive, and asymptomatically infected *superspreaders* identified during testing.

For this reason, data on absolute mortality from COVID-19 are more adequate for model verification than data on the number of infected. It is these data that were predominantly used to evaluate the re-wave criterion in the last section of our work.

Finally, note that the two-component model ignores the spatial heterogeneity of population density. This circumstance is important for countries with a large territory and an extensive system of regions and megalopolises (Russia, USA). For such countries, the epidemic acquires a multifocal character, and its spread to the entire territory of the country takes a longer time. Taking this circumstance into account requires considering the spatial distribution and corresponding modification of the model in the spirit of [2].

## Data Availability

Only open data sourses used

## ACKNOWLEDGEMENTS

I am deeply indebted to Prof. Mykola Iabluchanskiy and Dr. Vladimir Shlyachover for valuable remarks and discussions, and to Dr. Daniel Genin for help in use of Mathematika tools. I express my gratitude to Dr. Matteo Ferensby and to Dr. Giovani Vasconcelos for fruitful discussion of the results.

